# The role of Patient and Public Involvement (PPI) in pre-clinical spinal cord research: an interview study

**DOI:** 10.1101/2023.07.19.23292756

**Authors:** Pádraig Carroll, Adrian Dervan, Ciarán McCarthy, Ian Woods, Cliff Beirne, Geoff Harte, Dónal O’Flynn, Cian O’Connor, Tara McGuire, Liam M. Leahy, Javier Gutierrez Gonzalez, Martyna Stasiewicz, Jack Maughan, John Quinlan, Éimear Smith, Frank Moriarty, Fergal J. O’Brien, Michelle Flood

**Affiliations:** School of Pharmacy and Biomolecular Science, Royal College of Surgeons in Ireland University of Medicine and Health Sciences, Dublin, Ireland; Tissue Engineering Research Group (TERG), Department of Anatomy and Regenerative Medicine, Royal College of Surgeons in Ireland University of Medicine and Health Sciences, Dublin, Ireland; Advanced Materials and BioEngineering Research (AMBER) Centre, Trinity College Dublin (TCD) and RCSI, Dublin, Ireland; c/o Irish Rugy Football Union Charitable Trust, Dublin, Ireland; Faculty of Sports and Exercise Medicine (Royal College of Physicians in Ireland & RCSI), Dublin, Ireland; Tallaght University Hospital, Tallaght, Dublin, Ireland; National Rehabilitation Hospital, Dublin, Ireland; PPI Ignite Network, Galway, Ireland

**Keywords:** patient and public involvement, patient and public engagement, consumer involvement, public involvement in research, pre-clinical research, spinal cord injury

## Abstract

**Background:** Patient and public involvement in research (PPI) has many benefits including increasing relevance and impact. While using PPI in clinical research is now an established practice, the involvement of patients and the public in pre-clinical research, which takes place in a laboratory setting, has been less frequently described and presents specific challenges. This study aimed to explore the perspectives of seriously injured rugby players’ who live with a spinal cord injury on PPI in pre-clinical research.

**Methods:** Semi-structured interviews were conducted via telephone with 11 male seriously injured rugby players living with spinal cord injury on the island of Ireland. A purposive sampling approach was used to identify participants. Selected individuals were invited to take part via gatekeeper in a charitable organisation that supports seriously injured rugby players. Interviews were transcribed verbatim and analysed thematically.

**Findings:** Six themes were identified during analysis: ‘limited knowledge of PPI’, ‘connecting pre-clinical research with the day-to-day realities of spinal cord injury’, ‘making pre-clinical research accessible for non-scientific audiences’, ‘barriers to involvement include disinterest, accessibility issues, and fear of losing hope if results are negative’, ‘personal contact and dialogue facilitate PPI’, and ‘collaborating on dissemination builds trust in research.’

**Conclusion:** People affected by spinal cord injury in this study desire further involvement in pre-clinical spinal cord injury research through dialogue and contact with researchers. Sharing experiences of spinal cord injury can form the basis of PPI for pre-clinical spinal cord injury research.

## Introduction

Patient and public involvement (PPI) aims to ensure that research addresses the issues that are most important to patients and the public, and effects outcomes that are relevant and beneficial to them (1, 2). PPI has gained traction in clinical research (3) due to an improved understanding of its use and impact (2, 4, 5). In contrast to PPI in clinical research, focused on translating therapeutics from the laboratory to the bedside, the role of PPI in pre-clinical research is comparably less well-established (6). Pre-clinical research (referring to basic, fundamental, biomedical, translational, or laboratory-based research) involves research in the laboratory. This setting is removed from patients and the public’s day-to-day experiences of healthcare and may seem inaccessible or obscure when compared to clinical research (7). Some of the limited research indicates that PPI may offer benefits for pre-clinical research such as focusing research priorities to reduce research waste (8, 9) or incorporating patients’ perspectives to enhance the quality of research (10, 11). Integrating PPI into research has previously presented challenges for researchers, where a limited application has exposed PPI to criticisms of exclusivity and tokenism (12, 13). However, researchers and funding agencies have suggested that PPI be incorporated across all stages of the research cycle, both clinical and pre-clinical, to improve the relevance, quality, and validity of research (2, 14, 15).

Several studies have explored the experiences of members of the public involved in PPI (PPI contributors) (4, 5, 16, 17), who report an improved understanding of their condition, increased sense of self-worth, and feelings of empowerment and value (5, 17). As PPI is mostly understood in clinical research contexts, comparatively less is known about the experiences and perspectives of PPI contributors in pre-clinical research. PPI contributors who provided feedback on their experiences with pre-clinical PPI reported similar benefits such as feeling valued, building rapport with researchers to share experiences, and providing significant contributions to research activities (18, 19). However, some PPI contributors have also reported difficulty making meaningful contributions to pre-clinical research (18, 20). To overcome this challenge, regular communication and building relationships are seen have been suggested as enablers for pre-clinical PPI (20–22).

In spinal cord research, PPI has been used to develop patient-informed research priorities, which focus primarily on management, rehabilitation, and stem-cell-based therapeutics (23, 24). While there is an obvious role for PPI in management and rehabilitation research which already involves human participants, the potential for PPI in pre-clinical spinal cord research has not yet been realised. Spinal cord injury is one of the most traumatic injuries the human body can sustain (25), where surgical treatments remain limited (26). However, the development of new therapeutics continues, with much of the research at the pre-clinical stage (27–29). As the field develops, it is important for people with spinal cord injury to be involved in the pre-clinical research process, as they have been in clinical spinal cord research (24). Constructivist-based approaches may hold promise as a way of conducting PPI in pre-clinical research settings for spinal cord injury (22, 30).

Our PPI Advisory Panel noted the lack of information on the perspectives of people living with spinal cord injury when aiming to develop an evidence-informed PPI strategy to support a pre-clinical spinal cord repair project (29). The project in question is developing an advanced multifunctional biomaterial implant consisting of several technologies including stem cell and gene therapy-based approaches for spinal cord repair. The research is funded by a research partnership between the Irish Rugby Football Union (IRFU) Charitable Trust, and the Science Foundation Ireland (SFI) Advanced Materials and Bioengineering Research (AMBER) Centre conducted at RCSI University of Medicine and Health Sciences. Our panel comprises three seriously injured rugby players living with spinal cord injury, three clinicians with experience related to spinal cord interventions, surgery, and rehabilitation, pre-clinical researchers working in spinal cord research, and facilitators, and meets biannually to oversee the project, discuss findings, and collaborate on research (6). The team quickly identified that understanding patient and public perspectives on pre-clinical research would be useful in informing the evidence-based strategy and that while the PPI Advisory Panel incorporated the views of three seriously injured players, this might not be representative of the views of others. Therefore, this study aimed to explore seriously injured rugby players’ perspectives on PPI in pre-clinical spinal cord research.

The objectives of this interview-based study were:

1. to establish participants’ knowledge/experience of PPI
2. to explore how best to deliver/manage PPI
3. to identify barriers/enablers to participation in PPI

## Materials and Methods

### Study Design

This research employed a qualitative case study methodology with semi-structured interviews used for data collection (31). This approach allowed for an in-depth exploration of participants’ perspectives on pre-clinical PPI to achieve the study’s aim and objectives (32).

### Ethics

The Research Ethics Committee (REC) at RCSI provided ethical approval for the study (RCSI University of Medicine and Health Sciences REC Record ID: REC001697), followed by an amendment to the original ethics application to reflect an increase in the scope of the study. All participants provided written informed consent before participating in the study.

### Recruitment

Recruitment took place in June and July 2020. Purposive sampling was used to recruit participants with experience of serious rugby-related spinal cord injury (33). To obtain perspectives outside of those already on the advisory panel, the three seriously injured players who are members of the PPI Advisory Panel were not invited to participate in the interviews. The IRFU Charitable Trust office manager acted as a gatekeeper between the research team and prospective participants and shared the study invitation with seriously injured rugby players living with spinal cord injury, asking them to contact the research team if they were interested in participating. Participants were considered eligible if they had a rugby-related spinal cord injury and consented to participate in a 45-minute telephone interview. Those who contacted the study team were emailed a participant information leaflet containing details of the study and a consent form. PPI Advisory Panel members reviewed the information leaflets and consent forms before they were finalised to ensure they clearly outlined the remit of the study. Those who consented to participate were contacted to arrange a telephone interview. Participants were offered a €30 gift voucher in recognition of their time.

### Interviews

The interview topic guide (Appendix 1) was developed specifically for the study with input from members of the PPI Advisory Panel who provided feedback on the questions and terminology used in the interviews. The topic guide contained questions relating to participants’ knowledge of PPI, their perspectives on PPI in pre-clinical spinal cord research, and preferences for research dissemination. One pilot interview took place before commencing data collection. No changes to the topic guide were made based on the topic guide, and data collected in the pilot interview were not included in the analysis. Thereafter, telephone interviews were conducted between July and September 2020 by PC. PC is a male PhD student with a Master’s degree and formal training and experience in qualitative research. Interviews lasted 30-50 minutes. Interviews were audio recorded and transcribed verbatim by PC. No repeat interviews were carried out and participants were not asked to review their transcripts.

The concept of data saturation did not guide the number of interviews conducted in this research. Instead, several factors were considered to determine how many participants were required. As the aims of this study were narrow, aiming to explore PPI with people affected by spinal cord injury, fewer interviews would be required to achieve a high level of information power (34). Interviewing fewer participants is further justified by the specificity of the sample. In Ireland, there is a small population of people living with spinal cord injury, meaning a less extensive sample is required to illustrate their views. Finally, analysis in this study was conducted by case rather than cross-case, where participants answers are compared, therefore requiring fewer participants. According to the model proposed by Malterud et al (34), a study with narrow aims, specificity of participants, strong dialogue, and case analysis will typically require fewer participants.

### Analysis

Data were analysed using a thematic approach using a combination of deductive and inductive coding (35, 36). PC conducted the data analysis using NVivo12 (37), with regular feedback provided by MF throughout the process. Analysis was conducted using the framework set out by Braun and Clarke (38). First, transcripts were reread for familiarity; initial codes were then assigned using the main domains of the interview guide as a deductive framework. The framework consisted of three categories: (i) knowledge and views of PPI (ii) PPI for pre-clinical spinal cord research, and (iii) patient and public engagement (PPE). Next, an inductive approach was used to develop themes related to these categories. Themes were reviewed and refined by PC and MF before being finalised. A report was then produced. This study is reported according to the COnsolidated criteria for REporting Qualitative research (COREQ) Checklist (39).

### Patient and Public Involvement

This study was conducted as part of PC’s PhD research that aims to develop an evidence-informed PPI approach for pre-clinical spinal cord research with wider implications for pre-clinical research. Members of the PPI advisory panel were involved in the conceptualisation of this study, identification of the study aim, and informed the development of the interview protocol and topic guide by guiding wording and terminology (Appendix 1). The initial findings were also shared at a PPI advisory panel meeting, with feedback from the panel members incorporated into the discussion section of the manuscript. PPI panel members were also involved in revising this manuscript for important intellectual content.

## Results

### Participants

Eleven seriously injured players participated in the interviews out of the 33 individuals invited to participate, with a third of people contacted participating in the study (At the time the study was completed a total of 36 individuals were supported by the IRFU Charitable Trust (40) including the three members serving on the PPI Advisory Panel and who were deemed ineligible). All were male and living on the island of Ireland at the time of their interview. The findings are presented under the three categories as follows: (1) knowledge and views of PPI, (2) PPI for pre-clinical spinal cord research, and (3) engagement and dissemination. For the second and third categories, themes were developed.

### Findings

#### 1. Knowledge and Views of PPI

In the first part of the interview, participants were asked about their knowledge and views on PPI. PPI was defined for participants who were unaware of the concept.

#### 1.1 Limited knowledge of PPI

The majority of participants were unaware of PPI before taking part in the interview. None of the interviewees had any prior involvement in a PPI activity and only one participant had prior knowledge of PPI. The experienced interviewee viewed PPI as necessary for ensuring research is useful for people affected by it.

> *“I think it’s necessary* [PPI]*. If you’re not going to the person that’s going to use it* [the research], then it might have all the bells and whistles in the world but when you try to use it in the situation that it’s for, it’s not really useful.” **Participant 11**

To ensure that participants who did not know about PPI could answer the rest of the interview questions, a standard explanation was provided. When provided with a definition, participants indicated they felt it could be beneficial for research.

> “Well, I think it’s important. It offers perspective that the researcher might not otherwise be aware of.” **Participant 2**

#### 2. PPI for Pre-clinical Spinal Cord Research

The next part of the interview asked participants about PPI and the spinal cord repair project more specifically. Themes developed from their responses are outlined below.

#### 2.1 Connecting pre-clinical research with the day-to-day realities of spinal cord injury

Participants felt that there was a distance between pre-clinical spinal cord research taking place in a laboratory, and their daily experiences of living with a spinal cord injury. The majority spoke about the impact of their condition on routine activities, such as getting out of bed in the morning or taking public transport unassisted. Such experiences were seen as a form of knowledge, which could enhance researchers’ understanding of spinal cord injury and the priorities of those affected by it.

> “I do think that because given that people have sustained spinal cord injury. From their experience, they obviously are in a better position to offer their own experience of how this kind of thing has affected their lives and I think that the more they can offer to any research the better you know.” **Participant 6**
>
> “Obvious things, life changers, potential life changers that you’re not going to know or experts in the medical field aren’t going to know or engineering field aren’t going to know that exist for people with a spinal cord injury unless they talk to them.” **Participant 7**

Participants also spoke about their own priorities and how they may be different from those held by researchers. Participants noted that despite wanting to walk again, they held other priorities, developed over years of living with their condition. One also noted that researchers’ priorities may similarly evolve over time.

> “Some people often think that ‘oh sure he can’t walk’, but can’t walk? Jesus, I can think of 1001 things that I would like to have fixed before I’d be wanting to walk you know? You know that kind of thing, people think ‘Ah sure he can’t walk’, that’s the least of my worries to be honest.” **Participant 7**
>
> “Well, I suppose the priorities and areas that might make the biggest difference to their life. It would be easy with regards to getting people up and walking as the ultimate goal. Whereas you think that’s not necessarily the thing that should be the first goal. Now I see research into bladder function and the more personal elements of spinal cord injury that create a great problem for people. **Participant 5**

#### 2.2 Making pre-clinical research accessible for non-scientific audiences

When discussing pre-clinical spinal cord research, participants described difficulties understanding scientific language and terminology used by researchers. As pre-clinical research is often far removed from daily life, the language and terminology used are often specialised. Participants reported that the complex terminology often used by researchers prevents the non-scientific audience from understanding the significance of results, possibly leading to misunderstandings and unreasonably high expectations.

> *“It would be important they* [pre-clinical research findings] were explained to the public so they might be aware of the importance of it and just to simplify things that they would be obvious instead of having it too technical.” **Participant 7**

Presenting research for non-scientific audiences was seen as a responsibility for researchers. While some participants appreciated that this can be time-consuming, the majority felt it is important to minimise the potential for misunderstandings.

> “It’s almost like a translation from one thing to another and you don’t want research being misquoted indirectly…. the danger would be wrong information, more hope than should be necessary, misleading people unintentionally as well.” **Participant 8**
>
> “From my perspective, I think that the lab stage. So, the ability of the non-scientific community to understand, number 1, what you are doing. I think it’s probably important that somebody writes up an overview of the research that’s been done in layman’s terms and present it in a short format.” **Participant 2**

Participants felt there would be further benefits to disseminating research in a manner suitable for non-scientific audiences and could help educate the wider public on spinal cord research. One participant expressed that it may be a source of hope.

> “I think educating the public that this medical or research published has maintained that there may be long-term benefits of that sort could be helpful because ultimately they’d only ever get there if enough money, time, and brainpower were put into it.” **Participant 4**
>
> “Identifying key issues, key issues, key developments that have happened. I suppose basically to give people with spinal cord injuries a little bit of light or a little bit of hope. That I’ve seen in the world, living with a spinal cord injury every day is tough and there are always hiccups and hurdles, but just a small little glimmer of hope that could be spread around the community, any disability, it’s just like a little tonic.” **Participant 10**

#### 2.3 Barriers to involvement include disinterest, accessibility issues, and fear of losing hope if results are negative

Participants spoke about several barriers to PPI in pre-clinical spinal cord research. Some participants felt that many people living with spinal cord injuries had little to no interest in research. Pre-clinical research was also perceived to be slow moving, making it difficult for participants to stay interested throughout a research project.

> “The barriers would be I would think in my opinion, lack of interest. Just plain lack of interest.” **Participant 2**.
>
> “Trying to keep them fully involved in something that would be quite boring and tedious and long term and that could have its challenges too. That the experts could be at this for 50 years whereas this person mightn’t be able to give them that time to it you know.” **Participant 7**

Participants felt this barrier would be difficult to overcome, and that it would be difficult to persuade people without a research interest to become involved. One participant noted that many people with spinal cord injury prioritised other concerns related to their condition, over current research in spinal cord repair.

> “You would assume that the people with the most interest in this would be people with spinal cord injuries. Now in my experience, that’s not necessarily true. In other words, 90% of people with spinal cord injury have fig all interest in research, they have other fish to fry or there are bigger challenges in their lives or there could be any amount of reasons that it just doesn’t rock their boat and you just have to accept that.” **Participant 2**

Along with a disinterest, fear of losing hope if there were negative results was also reported as a barrier to PPI in pre-clinical spinal cord research. Some participants felt that becoming involved in research could confirm their belief that there is no cure for spinal cord injury.

Participants felt that managing expectations from pre-clinical research findings and communicating their significance clearly would be useful.

> “I suppose the main barrier is fear. Either ignoring the potential outcome and learning this from a PPI study. Fear of the outcome being positive fear of the outcome being negative or so many people sort of not wanting to know because they haven’t been affected in any way. They don’t know anyone who has been affected in any way.
>
> It’s almost as if, this is nothing to do with me so don’t tell me.” **Participant 6**
>
> “See, I would suspect a lot of people would view an idea of a repair project as ‘if you can repair the spinal cord, then everything will therefore come back and function.’ I doubt that would ever be the case. I could be proved wrong hopefully. But I think tempering people’s expectations and being firmly realistic in informing people what is and what are the outcomes would be most important and to that. Once you want people who are overly invested perhaps because they have the condition as a result of a spinal cord injury. They may want everything to come back to them depending on the amount of time that has passed from their injury. You know even if you were perfectly repaired for want of a better description, you’re going to be a much older human being at the end of it.” **Participant 3**

Finally, participants discussed the practical impact of spinal cord injury on taking part in PPI activities. Living with a spinal cord injury introduced physical barriers, which may not be readily apparent to pre-clinical researchers with a limited understanding of their condition.

Transport, health, and physical access to PPI meetings were seen as potential barriers to PPI. Participants highlighted that they relied on others in many aspects of their daily lives and that researchers should be flexible in organising PPI events. Technology, such as virtual meeting formats, was seen as an enabler of PPI.

> “Accessibility is the biggest thing really.” **Participant 5**.
>
> “You have to look at where there’s a start and an end for a person you know. You’re waiting for someone to get dressed in the morning and they don’t turn up. You know it’s game over straight away, you could be in bed all day. I know people who are waiting for the bus and the bus comes, and it’s full. You can’t hop on a taxi, you can’t hop on a bike. Yeah so sometimes it’s out of your control. If you’re willing, sometimes you need a little bit of patience for the other person.” **Participant 9**

#### 2.4 Personal contact and dialogue facilitate PPI

Participants indicated a preference for personal interaction and dialogue with pre-clinical researchers, rather than contributing via questionnaires or email. The majority of participants in this theme wanted to engage with the researchers directly and observe the research they were conducting on spinal cord repair.

> **“**I think they need to see the people, the real people that are behind the laboratory work.” **Participant 5**.
>
> “Being there, being present during some of the research rather than filling in a question-and-answer thing and sending it off to the research area you know? Rather than these surveys, you get from time to time. So being there and being present.” **Participant 8**

Furthermore, interaction and dialogue were seen as potential methods to build trust and clarity in research. Participants reported they would feel more valued from one-to-one interactions with researchers and more likely to have confidence in the research process as clarifications can be directly sought.

> “If you’ve got somebody who’s a researcher or something, and it’s as if you’re valued. So you would get an enormous amount of confidence in the whole thing if you were in a one-to-one with a researcher or scientist.” **Participant 8**
>
> “If I don’t understand what you say to me I can ask ‘what do you mean?’ Whereas if it’s in the form of a questionnaire, you’re maybe ‘I don’t know what they mean here and maybe I don’t give the information. They’re answering what they think and what they want to know rather than what you want to know and it’s not as beneficial as it could be.” **Participant 1**

#### 3. Engagement and dissemination

In the third stage of the interview, participants were asked about their priorities and preferences for engagement and the dissemination of research findings.

#### 3.1 Collaborating on dissemination builds trust in research

Most participants had an interest in spinal cord injury research and said they would like to see more pre-clinical spinal cord research disseminated to the spinal cord community. Several participants expressed interest in the IRFU/AMBER-funded pre-clinical spinal cord repair project conducted in RCSI and requested information on its nature.

> “I would always be interested in seeing what the most recent research is, what its objectives are, do they have a timeline? Where is it going? How does that fit into a bigger picture of research internationally? Are people collaborating? Are people talking to each other from different groups, countries, ideas? Yeah absolutely, it would be important to me to see for want of a better structure, any improvement in my situation.” **Participant 3**

When asked about engaging with research and dissemination, participants discussed the tendency for preliminary research findings to be sensationalised. Participants felt that news sites and social media tended to misinterpret and amplify pre-clinical research findings, and expressed a desire to see research disseminated in a more trustworthy manner.

> “I would be concerned that somebody who has not expertly involved might be slightly misinformed. Not deliberate, just their interpretation” **Participant 3**
>
> “I suppose it’s important that anyone who reads research in either of those forums again have the confidence that what they’re reading is relevant, is accurate, is not sort of scaremongering. That they can read and understand with a degree of confidence that what they’re reading is true and is accurate. It’s all about trust.” **Participant 6**

Therefore, many of the participants spoke of the potential benefits of collaborating with existing spinal cord injury patient organisations on disseminating pre-clinical research findings. Spinal cord organisations were seen as potential gatekeepers, ensuring that the research disseminated was relevant and accurate. Participants felt that pre-clinical researchers could collaborate with spinal cord injury patient organisations to disseminate research directly to those most likely to find it relevant.

> *“That’s where you know the likes of* [Spinal cord injury group]*, and others* [groups] get involved. You know where groups of spinally injured could get the information and feedback from their members. And could be passed on to other members”. **Participant 1**

## Discussion

### Summary of Findings

This study aimed to explore seriously injured rugby players’ perspectives on PPI in pre-clinical spinal cord research with specific objectives to (1) establish participants’ knowledge/experience of PPI, (2) explore how best to deliver/manage PPI and (3) identify barriers/enablers to participation. In this study, we asked participants to discuss the concept of PPI. That participants were unaware of PPI was not a surprising finding, as PPI (and other similar movements such as co-production and citizen science) is still a relatively new way of conducting research (1). When a definition for PPI was provided, participants were comfortable talking about PPI, as they reported that their lived experience could be of relevance to research. It is important that their perspectives are captured to encourage people without prior knowledge of involvement or engagement in pre-clinical research to take part in PPI. Representativeness of PPI contributors is a challenge for PPI (6), therefore exploring the perspectives of people without pre-existing awareness of PPI is useful. When discussing how PPI should be delivered, participants indicated that researchers should aim to bridge the gap between laboratory research and peoples’ daily experiences of spinal cord injury. However, the incremental nature of pre-clinical research and a fear of losing hope for a cure for their condition could deter people from becoming involved. Furthermore, they highlighted the practical impact of taking part in PPI activities when living with spinal cord injury, and that researchers should therefore be flexible. Finally, participants reported that contact and dialogue with researchers were enablers for PPI in pre-clinical spinal cord research, indicating that they would feel more valued by such interactions.

### Comparison with existing literature

#### 1. Knowledge and Views of PPI

It was not surprising that participants had little to no knowledge of PPI due to the limited amount that has taken place in pre-clinical research (6, 11). Upon hearing a definition of PPI, participants in our study were overwhelmingly positive about the involvement of people with spinal cord injury in pre-clinical research. Similar findings have been seen in the literature where PPI contributors report largely positive perspectives on the role and impact of PPI in pre-clinical research(18–20). Participants in our study held similar views to PPI contributors who took part in PPI. Namely, it adds an alternative perspective to those traditionally represented and is valuable to research (16). This also aligns with the literature on spinal cord research, where people living with spinal cord injury have indicated their desire to become more involved in research (30, 41, 42). A minority of pre-clinical researchers and PPI contributors have expressed scepticism about the contribution of PPI to laboratory research (18, 43), reporting that it would be difficult for PPI to make a meaningful impact. The perception that the public cannot influence pre-clinical research is understandable due to the apparent distances between a laboratory and real-world contexts. However, participants in our study felt their outlook was shaped by years of living with their condition, offering a valuable perspective not always represented in research.

#### 2. PPI for Pre-clinical Spinal Cord Research

Participants spoke about connecting non-patient-facing laboratory research with the daily realities of living with a serious condition. Previous studies have aimed to bridge this gap by facilitating meetings between patients and pre-clinical researchers (44, 45), or developing PPI advisory panels to collaborate throughout a research project (18, 46). Participants in our study discussed barriers to PPI in pre-clinical spinal cord research highlighting accessibility as a potential issue, which has also been seen in the spinal cord literature as a priority for spinal cord research (23). However, to our knowledge, outright disinterest due to more pressing psychological needs of the cohort and fear of losing hope has not been discussed as barriers to PPI.

The findings of this research study are also relevant to previous research on relationship building in pre-clinical PPI (18, 22, 46). Pre-clinical researchers and PPI contributors have previously expressed concerns over the potential for tokenism to emerge when PPI contributors have no clearly defined role in research (12, 13). Selective and tokenistic PPI approaches can be harmful to research, by presenting minority views as the majority (13), or through tokenistic application resulting in PPI contributors feeling unable to impact research (4). Whereas participants in our study propose that direct interaction and dialogue with researchers could facilitate a meaningful PPI process for pre-clinical spinal cord research. One participant highlighted this, stating: “*being there, being present during some of the research rather than filling in a question-and-answer thing and sending it off to the research area.”* Clear and regular communication between PPI contributors and researchers has been reported as an enabler for PPI in pre-clinical research (20, 22, 46). Participants in this study put similar emphasis on clear communication, familiar language, and accessible terminology as enablers for PPI in spinal cord research.

#### 3. Engagement and dissemination

Most of the participants in our study desired to see further dissemination of pre-clinical spinal cord research, though some concerns were raised about the language and terminology used by researchers. Previous research has suggested the general public hold misconceptions about pre-clinical research terminology (47). Public understanding of complex research topics depends on people’s personal understanding of scientific research, as the mechanisms behind research topics are rarely explained in current media at a depth to foster basic understanding (48). PPI may help bridge this gap and provide knowledge about laboratory research to the general public through dissemination activities (2). In previous research, PPI has been used to develop research materials to disseminate information about emerging biomedical research (49). This may hold promise for disseminating pre-clinical research findings relating to spinal cord injury.

### PPI Advisory Panel Perspectives

The themes developed during analysis and supporting quotes were presented to our PPI advisory panel (which constitutes the authorship team) for their feedback and comments at a panel meeting in 2022. All panel members noted that the findings were helpful in terms of planning the next steps for the Advisory Panel’s work and could help inform others undertaking PPI in pre-clinical spinal cord injury. Some specific comments made included one by one of the seriously injured players who responded to the comment about fear of negative results and lack of interest, explaining that in his experience, those living with spinal cord injury are usually advised by clinicians to ‘accept and adapt’ to their condition (50). He also felt that this may discourage those with spinal cord injury from being involved in PPI as it may appear to contradict this message. One pre-clinical researcher responded to the findings that concerns that fear of negative results was a barrier and clear communication about findings are needed, stating that pre-clinical research makes incremental progress, and therefore expectations should be carefully managed as ‘even the lowest hanging fruit is quite high’. Finally, one clinician considered theme 2.2 *Making pre-clinical research accessible for non-scientific audiences* was relevant to clinicians, and that accessibility went beyond just PPI, where the language used by pre-clinical researchers often prevented clinicians from fully understanding pre-clinical research findings. These perspectives were recorded for incorporation into our research and future planning.

### Limitations

Before this study, the interviewed participants had limited experience and knowledge of PPI and pre-clinical research. While PPI and pre-clinical research were both defined and explained at the beginning of the interview, it is possible that some participants were still not fully clear on the full scope of PPI in pre-clinical research. Despite this, they represented potential future PPI contributors, so their perspectives were valid. As this research is conducted in the context of rugby-related spinal cord injury, findings may not be representative of the entire spinal cord injury community. Participants in this study participated voluntarily and were all male. While all eligible seriously injured rugby players of the associated charity were contacted for recruitment, those who chose not to participate may hold perspectives not represented in the findings of this research.

### Implications

Through analysis of data and PPI advisory panel meetings, we identify several considerations for PPI in pre-clinical spinal cord research. While the findings and implications of this research are conducted in the context of pre-clinical spinal cord research, they are also relevant for PPI in pre-clinical research. Namely, the findings of this study suggest that PPI should focus on connecting research with the day-to-day realities of spinal cord injury.

Considerations for PPI in pre-clinical spinal cord research are outlined below:

1. Make clear the links between pre-clinical spinal cord research and its real-world application to facilitate PPI.
2. Use terminology and language familiar to non-scientific audiences to ensure that pre-clinical spinal cord research is accessible.
3. Focuses on optimising interest, addressing accessibility concerns, and clearly communicating the meaning of research findings.
4. Contact and dialogue between researchers and people affected by spinal cord injury can form a basis for PPI in pre-clinical spinal cord research activities.
5. Researchers should involve people affected by spinal cord injury in disseminating research findings to build trust and increase impact.

## Conclusions

People affected by rugby-related spinal cord injury want further involvement in pre-clinical research, but in order to do so several mechanisms need to be in place in order for meaningful interaction and potential collaboration to occur. This can be achieved by reducing the distance between laboratory-based science and the day-to-day experiences of people living with spinal cord injuries. Sharing experiences of spinal cord injury and disseminating research for non-scientific audiences may enable PPI. However, dispelling the lack of research interest, fear of negative research outcomes, and accessibility are challenges to overcome for PPI in pre-clinical spinal cord research.

## Data Availability

All data produced in the present study are available upon reasonable request to the authors

## Acknowledgements

This study is conducted as part of a research collaboration funded by the Irish Rugby Football Union Charitable Trust (IRFU CT), Science Foundation Ireland Advanced Materials and BioEngineering Research Centre (SFI AMBER) and conducted by Tissue Engineering Research Group (TERG) at the Royal College of Surgeons in Ireland (RCSI) University of Medicine and Health Sciences. Pádraig Carroll is also a recipient of a Clement Archer Postgraduate Scholarship from the RCSI School of Pharmacy and Biomolecular Sciences.

## Appendix 1

### Topic Guide

Interviewer to welcome participant to the session, remind them that it is intended to be an informal discussion and their honest opinions (good or bad) are welcome.

Interviewer to invite questions, ensure the consent form has been completed, remind participant of confidentiality of data, and seek permission to start recording.

**Part A: Background/Personal Views**

1. Have you ever heard the term ‘patient and public involvement in research’ or ‘PPI’? What do you think it might mean? Note 1: The interviewer will then tell the participant that ‘PPI’ will be used in the interview (this is suggested based on pilot participants’ feedback). Note 2: If the participant does not know what it is or gives an incomplete/inaccurate answer, the INVOLVE definition will be provided: *INVOLVE, a group that develop PPI policy, defines public involvement in research as research being carried out ‘with’ or ‘by’ members of the public rather than ‘to’, ‘about’ or ‘for’ them. This includes, for example:*

- working with research funders to prioritise research;
- offering advice as members of a project steering group;
- commenting on and developing research materials;
- undertaking interviews with research participants.

1. What is your opinion about PPI in research [based on this definition]?
2. Have you ever participated in a PPI activity? E.g. training, a workshop etc.?
  - How did you come to be involved?
  - What was it about?
  - What did you think about it?

1. In your experience as a person affected by spinal cord injury, would you consider PPI in research important to you?
2. Why/why not?
3. How important is it for you to personally participate in patient and public involvement activities and why?

**Part B: The Spinal Cord Repair Project**

As explained in the participant information leaflet, the project aims to find new ways to repair spinal cord injury. This research is being conducted by research scientists in a laboratory setting, for example, on animal models or with human cells.

1. What should the goals of PPI for this research be from your perspective and why?
2. Some examples of times in research where patient and public involvement is used, include the following, what do you think is good/bad about each in the context of the spinal cord repair project and why?
3. ‘Priority Setting’ – e.g. helping to identify research needs, ensure research is relevant to those affected by particular conditions, highlight potential future new directions
4. ‘Management/oversight activities’ – e.g. sitting on project steering groups, or acting as a member of an advisory panel
5. ‘Application and Design’ – e.g. review and write ‘lay material’ such as articles or blogs, design material for people affected by the condition
6. ‘Dissemination/communicating research findings’ – e.g. identifying key findings to share with the public, helping to plan outreach/sharing activities for the public.
7. In what other ways might people affected by spinal cord injury be involved in your opinion?
8. In your opinion, what barriers might exist to involving people affected by spinal cord injury in PPI activities and how might they be overcome?
9. What would make it easier to involve people affected by spinal cord injury in PPI and why?

**C. Patient and Public Engagement**

‘Patient and Public Engagement’ or PPE is another way that people are included in research activities, often to allow sharing of information about completed research activities such as findings or results.

1. Is this important to you as a person affected by spinal cord injury? Why?
2. The following methods are commonly used to engage people affected by particular conditions with research.

What do you think is good or bad about the following options in terms of the spinal cord repair project and why?

- Social media (e.g. Twitter or blogging)
- Talks held in a college/university for the public
- Exhibitions at museums/discovery centres or festivals
- Science talks held in cafés or bars
- Media broadcasts (television or radio)
- Research-buddies (having a 1:1 relationship with a specific researcher)
- Are there any other ways you think might be good ways to involve/engage people in spinal cord research?

**D. Closing**

1. Is there anything that we have not discussed that you would like to mention now?
2. What questions do you have for me?

Thank you for your time.

## Notes

### Competing Interest Statement

The authors have declared no competing interest.

### Author Declarations

Ethical approval was granted by the Research Ethics Committee (REC) at the Royal College of Surgeons in Ireland, University of Medicine and Health Sciences - Reference number: REC001697

## References

1. Russell J, Greenhalgh T, Taylor M. Patient and public involvement in NIHR research 2006– 2019: policy intentions, progress and themes. National Institute for Health Research: Oxford, UK. 2019.

2. Natioanl Institute for Health Research. Briefing notes for researchers - public involvement in NHS, health and social care research 2022 [Available from: http://www.nihr.ac.uk]

3. Richards T. Patient and public involvement in research goes global. BMJ Patient Perspect. 2017.

4. Brett J, Staniszewska S, Mockford C, Herron-Marx S, Hughes J, Tysall C, et al. A systematic review of the impact of patient and public involvement on service users, researchers and communities. The Patient-Patient-Centered Outcomes Research. 2014;7(4):387–95.

5. Brett J, Staniszewska S, Mockford C, Herron-Marx S, Hughes J, Tysall C, et al. Mapping the impact of patient and public involvement on health and social care research: a systematic review. Health expectations. 2014;17(5):637–50.

6. Carroll P, Dervan A, Maher A, McCarthy C, Woods I, Kavanagh R, et al. Applying Patient and Public Involvement in preclinical research: A co-created scoping review. Health Expectations. 2022;25(6):2680–99.

7. Allum N, Sturgis P, Tabourazi D, Brunton-Smith I. Science knowledge and attitudes across cultures: A meta-analysis. Public understanding of science. 2008;17(1):35–54.

8. Macleod MR, Michie S, Roberts I, Dirnagl U, Chalmers I, Ioannidis JP, et al. Biomedical research: increasing value, reducing waste. The Lancet. 2014;383(9912):101–4.

9. Minogue V, Cooke M, Donskoy A-L, Vicary P, Wells B. Patient and public involvement in reducing health and care research waste. Research involvement and engagement. 2018;4(1):1–8.

10. Caron-Flinterman JF, Broerse JE, Bunders JF. The experiential knowledge of patients: a new resource for biomedical research? Social science & medicine. 2005;60(11):2575–84.

11. Fox G, Fergusson DA, Daham Z, Youssef M, Foster M, Poole E, et al. Patient engagement in preclinical laboratory research: A scoping review. EBioMedicine. 2021;70:103484.

12. Ocloo J, Matthews R. From tokenism to empowerment: progressing patient and public involvement in healthcare improvement. BMJ quality & safety. 2016;25(8):626–32.

13. Russell G, Starr S, Elphick C, Rodogno R, Singh I. Selective patient and public involvement: The promise and perils of pharmaceutical intervention for autism. Health Expectations. 2018;21(2):466–73.

14. de Wit MP, Berlo SE, Aanerud G-J, Aletaha D, Bijlsma J, Croucher L, et al. European League Against Rheumatism recommendations for the inclusion of patient representatives in scientific projects. Annals of the rheumatic diseases. 2011;70(5):722–6.

15. Selby JV, Beal AC, Frank L. The Patient-Centered Outcomes Research Institute (PCORI) national priorities for research and initial research agenda. Jama. 2012;307(15):1583–4.

16. Crocker JC, Boylan AM, Bostock J, Locock L. Is it worth it? Patient and public views on the impact of their involvement in health research and its assessment: a UK-based qualitative interview study. Health Expectations. 2017;20(3):519–28.

17. Wilson P, Mathie E, Keenan J, McNeilly E, Goodman C, Howe A, et al. ReseArch with Patient and Public invOlvement: a realisT evaluation: the RAPPORT study. Health services and delivery research. 2015.

18. Birch R, Simons G, Wähämaa H, McGrath CM, Johansson EC, Skingle D, et al. Development and formative evaluation of patient research partner involvement in a multi-disciplinary European translational research project. Research involvement and engagement. 2020;6(1):1–14.

19. Bradshaw E, Whale K, Burston A, Wylde V, Gooberman-Hill R. Value, transparency, and inclusion: A values-based study of patient involvement in musculoskeletal research. Plos one. 2021;16(12):e0260617.

20. de Souza S, Johansson EC, Karlfeldt S, Raza K, Williams R. Patient and public involvement in an international rheumatology translational research project: an evaluation. BMC rheumatology. 2022;6(1):1–16.

21. Costello W, Dorris E. Laying the groundwork: Building relationships for public and patient involvement in pre-clinical paediatric research. Health Expectations. 2020;23(1):96–105.

22. Baart IL, Abma TA. Patient participation in fundamental psychiatric genomics research: a Dutch case study. Health Expectations. 2011;14(3):240–9.

23. Hammell KRW. Spinal cord injury rehabilitation research: patient priorities, current deficiencies and potential directions. Disability and rehabilitation. 2010;32(14):1209–18.

24. Van Middendorp J, Allison H, Ahuja S, Bracher D, Dyson C, Fairbank J, et al. Top ten research priorities for spinal cord injury: the methodology and results of a British priority setting partnership. Spinal Cord. 2016;54(5):341–6.

25. Jazayeri SB, Beygi S, Shokraneh F, Hagen EM, Rahimi-Movaghar V. Incidence of traumatic spinal cord injury worldwide: a systematic review. European spine journal. 2015;24:905–18.

26. Haldrup M, Schwartz OS, Kasch H, Rasmussen MM. Early decompressive surgery in patients with traumatic spinal cord injury improves neurological outcome. Acta Neurochirurgica. 2019;161:2223–8.

27. Courtine G, Sofroniew MV. Spinal cord repair: advances in biology and technology. Nature medicine. 2019;25(6):898–908.

28. Bradbury EJ, Burnside ER. Moving beyond the glial scar for spinal cord repair. Nature communications. 2019;10(1):3879.

29. Woods I, O’Connor C, Frugoli L, Kerr S, Gutierrez Gonzalez J, Stasiewicz M, et al. Biomimetic Scaffolds for Spinal Cord Applications Exhibit Stiffness-Dependent Immunomodulatory and Neurotrophic Characteristics. Advanced Healthcare Materials. 2022;11(3):2101663.

30. Abma TA. Patients as partners in a health research agenda setting: the feasibility of a participatory methodology. Evaluation & the health professions. 2006;29(4):424–39.

31. Cousin G. Researching learning in higher education: An introduction to contemporary methods and approaches: Routledge; 2009.

32. Cheek C, Hays R, Smith J, Allen P. Improving case study research in medical education: a systematised review. Medical education. 2018;52(5):480–7.

33. Etikan I, Musa SA, Alkassim RS. Comparison of convenience sampling and purposive sampling. American journal of theoretical and applied statistics. 2016;5(1):1–4.

34. Malterud K, Siersma VD, Guassora AD. Sample size in qualitative interview studies: guided by information power. Qualitative health research. 2016;26(13):1753–60.

35. Fereday J, Muir-Cochrane E. Demonstrating rigor using thematic analysis: A hybrid approach of inductive and deductive coding and theme development. International journal of qualitative methods. 2006;5(1):80–92.

36. Clarke V, Braun V, Hayfield N. Thematic analysis. Qualitative psychology: A practical guide to research methods. 2015;222:248.

37. QSR international Pty Ltd. (2020) NVivo (released in March 2020) [Available from: https://www.qsrinternational.com].

38. Braun V, Clarke V. Is thematic analysis used well in health psychology? A critical review of published research, with recommendations for quality practice and reporting. Health Psychology Review. 2023:1–24.

39. Tong A, Sainsbury P, Craig J. Consolidated criteria for reporting qualitative research (COREQ): a 32-item checklist for interviews and focus groups. International journal for quality in health care. 2007;19(6):349–57.

40. Irish Rugby Football Union Charitable Trust. Our Purpose 2023 [cited 2023 03/02/2023]. Available from: https://irfucharitabletrust.com/about-us/.

41. Abma TA. Patient participation in health research: research with and for people with spinal cord injuries. Qualitative health research. 2005;15(10):1310–28.

42. Abma TA, Pittens CA, Visse M, Elberse JE, Broerse JE. Patient involvement in research programming and implementation: a responsive evaluation of the dialogue model for research agenda setting. Health Expectations. 2015;18(6):2449–64.

43. Gibson A, Kok M, Evans D, Grier S, MacGowan A. Challenges and opportunities for involving patients and the public in acute antimicrobial medicine development research: an interview study. BMJ open. 2019;9(4):e024918.

44. Chalasani M, Vaidya P, Mullin T. Enhancing the incorporation of the patient’s voice in drug development and evaluation. Research Involvement and Engagement. 2018;4(1):1–6.

45. Elberse JE, Pittens CACM, de Cock Buning T, Broerse JEW. Patient involvement in a scientific advisory process: setting the research agenda for medical products. Health Policy. 2012;107(2-3):231–42.

46. Supple D, Roberts A, Hudson V, Masefield S, Fitch N, Rahmen M, et al. From tokenism to meaningful engagement: best practices in patient involvement in an EU project. Research Involvement and Engagement. 2015;1(1):1–9.

47. Lanie AD, Jayaratne TE, Sheldon JP, Kardia SL, Anderson ES, Feldbaum M, et al. Exploring the public understanding of basic genetic concepts. Journal of genetic counseling. 2004;13(4):305–20.

48. Miller JD. Public understanding of, and attitudes toward, scientific research: What we know and what we need to know. Public understanding of science. 2004;13(3):273–94.

49. Diamond J, Jee B, Matuk C, McQuillan J, Spiegel AN, Uttal D. Museum monsters and victorious viruses: improving public understanding of emerging biomedical research. Curator: The museum journal. 2015;58(3):299–311.

50. Aaby A, Ravn SL, Kasch H, Andersen TE. The associations of acceptance with quality of life and mental health following spinal cord injury: a systematic review. Spinal Cord. 2020;58(2):130–48.

